# Pushing beyond specifications: Evaluation of linearity and clinical performance of a fully automated SARS-CoV-2 RT-PCR assay for reliable quantification in blood and other materials outside recommendations

**DOI:** 10.1101/2020.05.28.20115469

**Authors:** Dominik Nörz, André Frontzek, Ulrich Eigner, Lisa Oestereich, Nicole Fischer, Martin Aepfelbacher, Susanne Pfefferle, Marc Lütgehetmann

## Abstract

1

**Background:** The ongoing SARS-CoV-2 pandemic presents a unique challenge to diagnostic laboratories. There are preliminary studies correlating qRT-PCR results from different materials to clinical outcomes, yet, comparability is limited due to the plethora of different assays used for diagnostics. In this study we evaluate clinical performance and linear range for the SARS-CoV-2 IVD (cobas6800/8800 system, a fully automated sample-to-result platform) in different clinically relevant matrix materials outside official specifications.

**Methods:** Assay performance was assessed in human plasma, BAL/BL and transport medium following chemical inactivation. For analytical evaluation, respective matrix materials were spiked with SARS-CoV-2 RNA in ten-fold dilution series. The efficacy of chemical inactivation by guanidine hydrochloride solution was confirmed in cell culture infectivity experiments. For correlation, a total of 235 predetermined clinical samples including respiratory swabs, plasma and BAL/BL were subjected to the SARS-CoV-2 IVD test and results were compared.

**Results:** The SARS-CoV-2 IVD showed excellent linearity over five to seven log steps depending on matrix material. Chemical inactivation resulted in a reduction in plaque forming units of at least 3.5 log steps, while having no significant impact on assay performance. Inter-run consistency from three different testing sites demonstrated excellent comparability of RT-PCR results (maximum deviation was 1.53 CT). Clinical evaluation for respiratory swabs showed very good agreement with the comparator assay (Positive agreement 95.7%, negative agreement 98.9%).

**Conclusion:** The SARS-CoV-2 IVD test for the cobas6800/8800 systems offers excellent linear range and inter-run consistency for quantification of SARS-CoV-2 RNA in different matrices outside official specifications.

**Highlights:**

- Effective reduction of SARS-CoV-2 infectivity by chemical inactivation without affecting assay performance.
- SARS-CoV-2 IVD for the cobas 6800/8800 is linear over up to seven log steps in different materials including human plasma.
- Minimal variance of CT values between testing sites indicates high comparability of quantification results.

## 2 Introduction

The novel coronavirus SARS-CoV-2 has been subject of extensive study since its emergence in late December 2019 (1, 2). Diagnostic RT-PCR was quickly determined as the gold standard for detecting the new pathogen in patients, in large parts due to the rapid dissemination of complete virus sequences from the assumed origin of the outbreak in China and consecutive publication of specific PCR assays (3, 4). Besides merely confirming the diagnosis, there exists evidence for a correlation between viral loads and clinical outcomes for various respiratory viruses, including Influenza and the original SARS-CoV (5, 6). In the case of SARS-CoV-2 there has already been a flurry of publications describing viral load dynamics in different clinical specimens (7, 8). It can be assumed that this topic will further grow in relevance in the coming months.

Reliable quantification of RT-PCR signals is highly dependent on assay specifications and reference materials (9). In the absence of an international standard to serve as reference, quantification of SARS-CoV-2 RNA is ultimately based on a variety of different methods and standards used in the individual labs (8, 10), resulting in inherently poor comparability and reproducibility between testing sites. However, the increasing availability of commercial SARS-CoV-2 RT-PCR kits for widely used fully automated systems offers the opportunity to generate highly consistent PCR results which can then be used for quantification.

The cobas6800 system is a fully automated sample-to-result high-throughput RT-PCR platform, capable of performing 384 tests in an 8 hour shift and requiring minimal hands-on time (11). Until very recently, laboratory developed tests (LDT) using the open mode (utility channel) had to be employed to use the system for SARS-CoV-2 testing (10). However, Roche has since received “Emergency use authorization” by the FDA for their own SARS-CoV-2 IVD assay and have made the test available commercially (12). This provides a common ground across different testing sites for quantification of RT-PCR data, especially when taking the remarkable inter-run consistency of the instrument into account (13).

The aim of this study was to provide clinical evaluation of the new SARS-CoV-2 IVD assay by Roche and further validate clinical specimen types outside specifications for use in clinical studies. In this context, efficiency and linear range was to be determined in different clinical materials. Lastly, comparability of results was to be evaluated using a multicenter approach for inter-run variability.

## 3 Materials and Methods

### 3.1 Cell culture and virus quantification

Vero cells (ATCC CCL 81) were propagated in DMEM containing 10 % FCS, 1% Penicillin/Streptomycin, 1 % L-Glutamine, (200 mM), 1 % Sodium pyruvate and 1 % non-essential amino acids (all Gibco/ Thermo Fisher, Waltham, USA). A SARS-CoV-2 isolate was rescued from an oropharyngeal swab as described previously (Pfefferle et al., manuscript submitted). For virus quantification, plaque-assays were performed as follows: A tenfold serial dilution series of the virus containing solution was incubated on Vero cells seeded in 24-well plates (500 µl per well). After adsorption at 37°C for 1h, cells were washed once with PBS and overlaid with DMEM containing 1% methylcellulose to prevent virus spreading within the supernatant. After 5 days of incubation at 37°C, cells were fixed in 4% formaldehyde (30 min at room temperature), washed once with H_2_O and stained with crystal violet solution. Virus titers in initial (stock) solutions were assessed by counting plaques.

### 3.2 Assessment of the virus inactivation capability of 40% guanidine hydrochloride

In order to determine the virus inactivation capability of the Roche PCR Media Kit (≤ 40% guanidine hydrochloride in Tris-HCl), 100 µl of the SARS-CoV-2 stock solution (with an estimated 3×10^7^ PFU/ml) were diluted 1:10 in either UTM or E-swab medium (modified Amies medium in Tris-HCl). 500 µl of each dilution were supplemented 1:1 with Roche PCR-Media, the remaining 500µl were supplemented 1:1 with either UTM or E-swab medium. After 30 min of incubation at room temperature, ten-fold serial dilution series of each mixture were used for plaque assays as described above.

### 3.3 Cobas6800 SARS-CoV-2 IVD Test setup

The SARS-CoV-2 IVD dual-target test for the cobas6800/8800 system was obtained from Roche and used according to instructions issued by the manufacturer. Target-1 (RdRp gene) and Target-2 (E gene) properties were analysed separately, though the entire assay was deemed positive as long as one Target returned a positive result. Apart from loading the ready-to-use SARS-CoV-2 IVD-test cartridges onto the device, there are no manual steps required in the assay setup.

### 3.4 Assessing linearity in different materials

For determining linear range in different matrix materials, an initial 1:20 dilution of cell culture supernatant containing SARS-CoV-2 (isolate HH-1) was prepared using normal human plasma (SARS-CoV-2 RNA negative) as diluent. For BAL/BL, clinical specimens previously tested PCR-negative for SARS-CoV-2 were pooled to serve as matrix material. The initial stock was used to generate tenfold dilution series, which were subjected to the SARS-CoV-2 IVD test in 8 repeats for each step.

### 3.5 Comparing clinical samples

All tested clinical samples were predetermined positive or negative in routine diagnostics using the SARS-CoV-2 UCT (utility channel test) on the cobas6800 system (10). Oropharyngeal swabs or nasopharyngeal swabs were collected using eSwab collection kits (Copan, Italy). All samples were pre-treated by adding 1:1 “cobas PCR media” (Roche, ≤ 40% guanidine hydrochloride in Tris-HCL buffer) for safe handling. To obtain EDTA-plasma, EDTA-blood samples were separated by centrifugation and transferred into fresh sample tubes.

A total of 180 respiratory swab samples, 43 positive plasma samples and 12 positive BAL/BL samples were retrieved from storage at –20°C (< 1 month) and subjected to the SARS-CoV-2 IVD test. CT values of Target-1 (RdRp gene) and Target-2 (E gene) were compared to the prior result of the comparator assay (targeting the viral E gene).

This work was conducted in accordance with §12 of the Hamburg hospital law (§12 HmbKHG). The use of anonymized samples was approved by the ethics committee, Freie und Hansestadt Hamburg, PV5626.

### 3.6 Inter-run variability and comparability of results

Inter-run variability was compared between a total of four different instruments, located at 3 different testing sites. Triplicates of a solution containing diluted SARS-CoV-2 infected cell culture supernatant were prepared as described above. Concentrations of SARS-CoV-2 RNA were estimated to be within linear range of the assay. Spiked sample sets were sent to “Labor Limbach”, Heidelberg and “Labor Stein”, Moenchengladbach for analysis with the SARS-CoV-2 IVD test using their own cobas6800 instruments. Ct values were reported and compared. The total number of measurements for each series was 11.

## 4 Results

### 4.1 Supplementing swab medium with guanidine hydrochloride solution results in significant reduction of plaque formation

To confirm effective neutralization of infectious virus in clinical specimens, cell culture supernatants containing high concentrations of SARS-CoV-2 (HH-1) were initially diluted (1:20) in UTM and eSwab medium, with and without adding 1:1 with Roche cobas PCR media (≤ 40% guanidine hydrochloride in Tris-HCL buffer) to the mix. Stocks treated with guanidine hydrochloride solution showed a reduction in plaque forming units of at least 3.5 log steps compared to control (*figure 1*). No significant differences were observed when comparing PCR efficiency in UTM to UTM or eSwab medium supplemented with guanidine hydrochloride solution (*figure 2, Supplement 1*).

**Figure 1:**
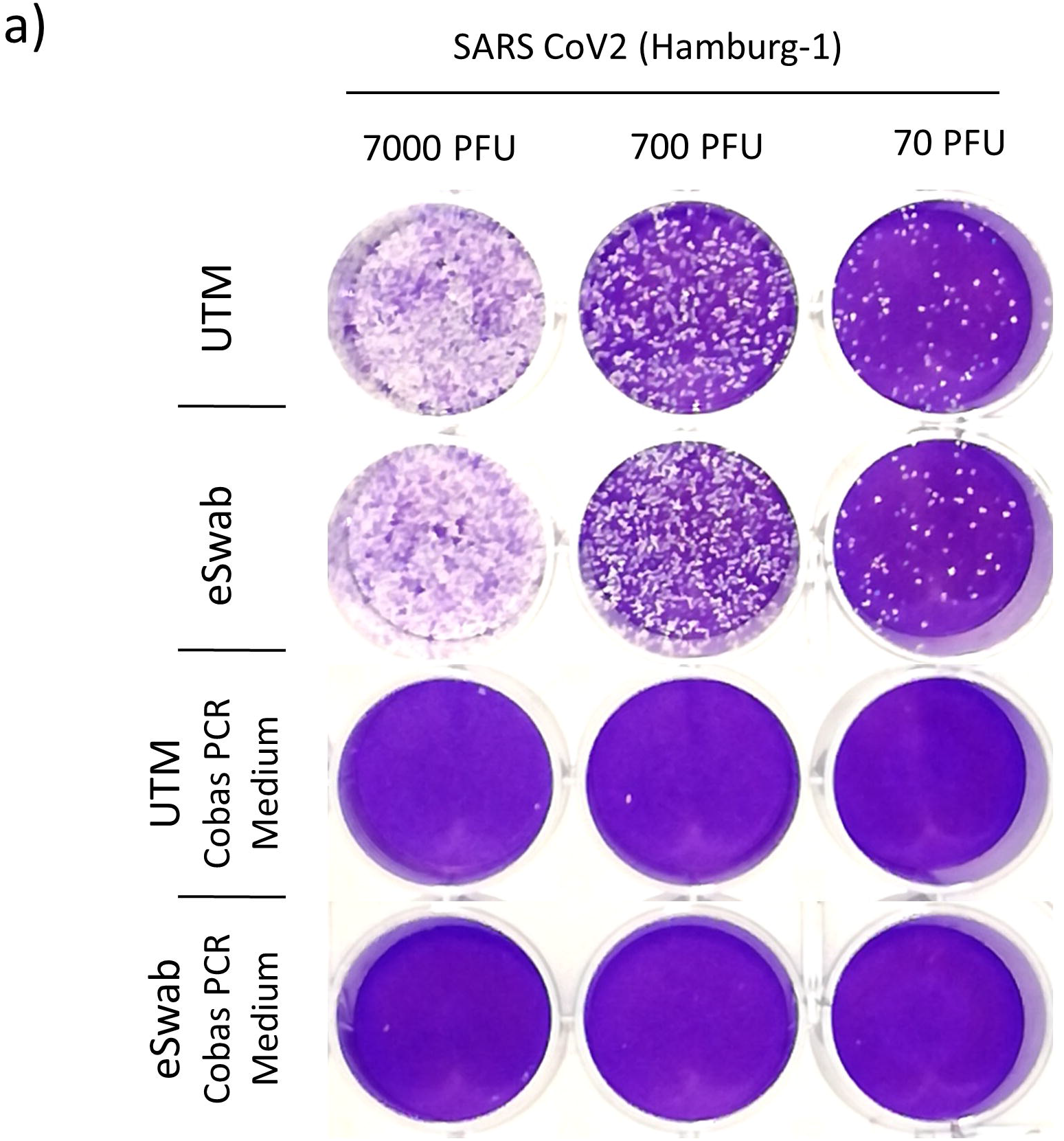
Reduction of plaque forming units (PFU) by guanidine hydrochloride solution. A stock solution of cell culture supernatant containing SARS-CoV-2 (HH-1) was diluted 1:20 in UTM or eSwab medium alone or supplemented 1:1 with cobas PCR Media. A 10-fold dilution series was prepared and 200µL added to Vero cells grown to fluency in a 24 well plate.

**Figure 2:**
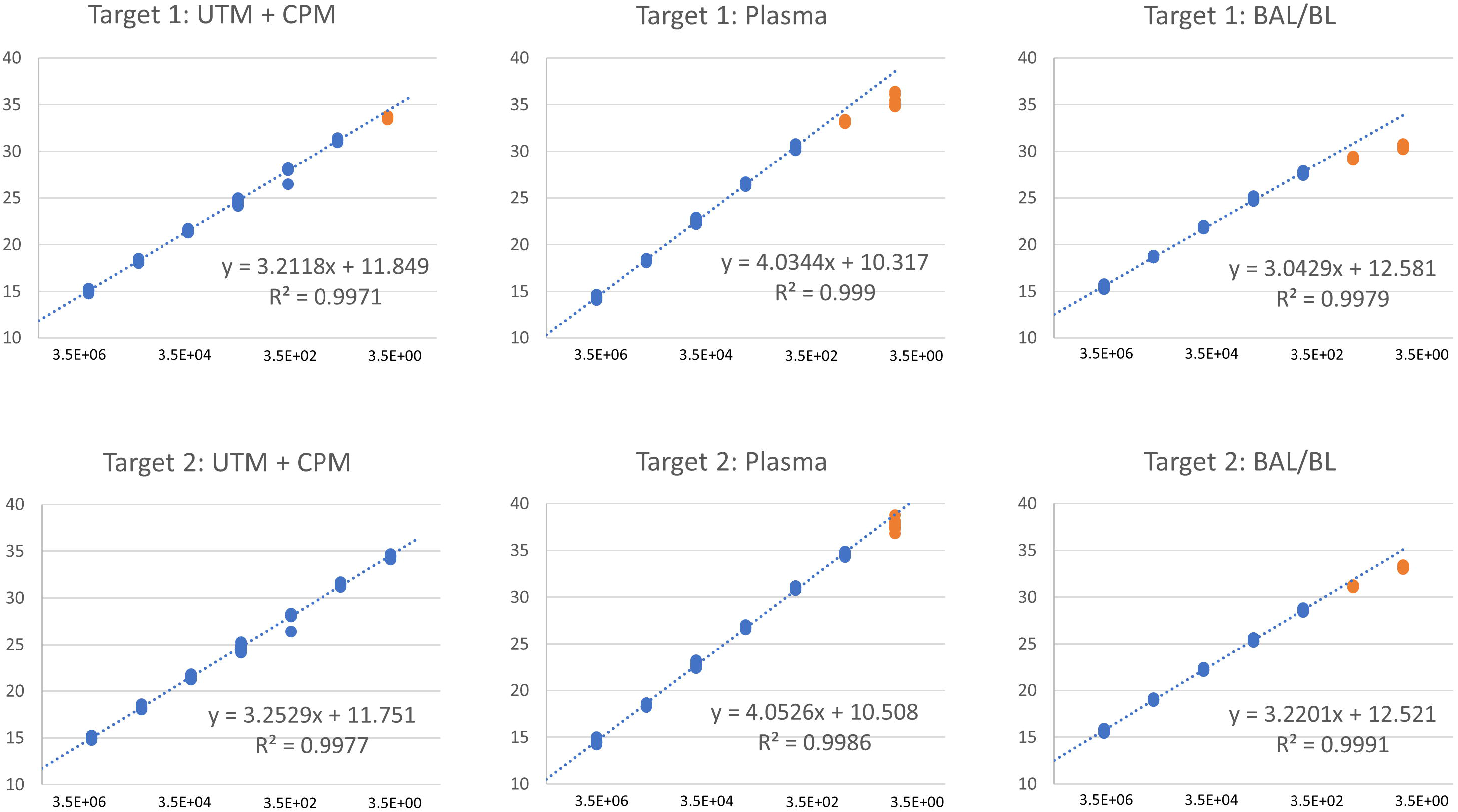
Linear range of Target-1 and Target-2 in different matrix materials. A stock solution of cell culture supernatant containing SARS-CoV-2 (HH-1) was used to prepare a 10-fold dilution series within indicated materials. A total of 8 repeats was tested per dilution step. Blue dots: measurements within linear range of Target-2, considered for trendline and correlation. Orange dots: measurements outside linear range, not considered for trendline and correlation. CPM: cobas PCR media (≤ 40% guanidine hydrochloride in Tris-HCl).

### 4.2 Linear range for different matrix materials using cell culture stocks

Linear range of the assay was assessed in a variety of matrices, including swab medium with and without chemical inactivation, human plasma and BAL/BL. Both Targets exhibited slopes in the range of 3 to 3.3 CT in all tested materials except plasma, indicating very good efficiency (*figure 2, supplement 1*). Slopes were markedly higher in plasma, approximately 4 CT for both Targets. The same behavior was observed with the SARS-CoV-2 UCT (*Supplement 2*).

Both assays demonstrated excellent linearity over a range of 5 – 7 log steps (7 log in UTM incl. guanidine hydrochloride, 6 log in plasma, 5 log in BAL/BL), with the Target-2 assay performing slightly better and being highly linear up to a CT of 35.

### 4.3 SARS-CoV-2 IVD results in clinical samples within linear range show excellent correlation with the comparator assay

For clinical validation, a set of predetermined samples was subjected to testing with the SARS-CoV-2 IVD and results were compared to the comparator assay. The SARS-CoV-2 IVD result was reported as “positive” as long as one of the two Targets returned a positive result. A total of 180 respiratory swabs (predetermined positive: 93; predetermined negative: 87) tested with the assay resulted in robust positive and negative agreement of 95.7 % and 98.9 % respectively (*figure 3a, table 1*).

**Figure 3:**
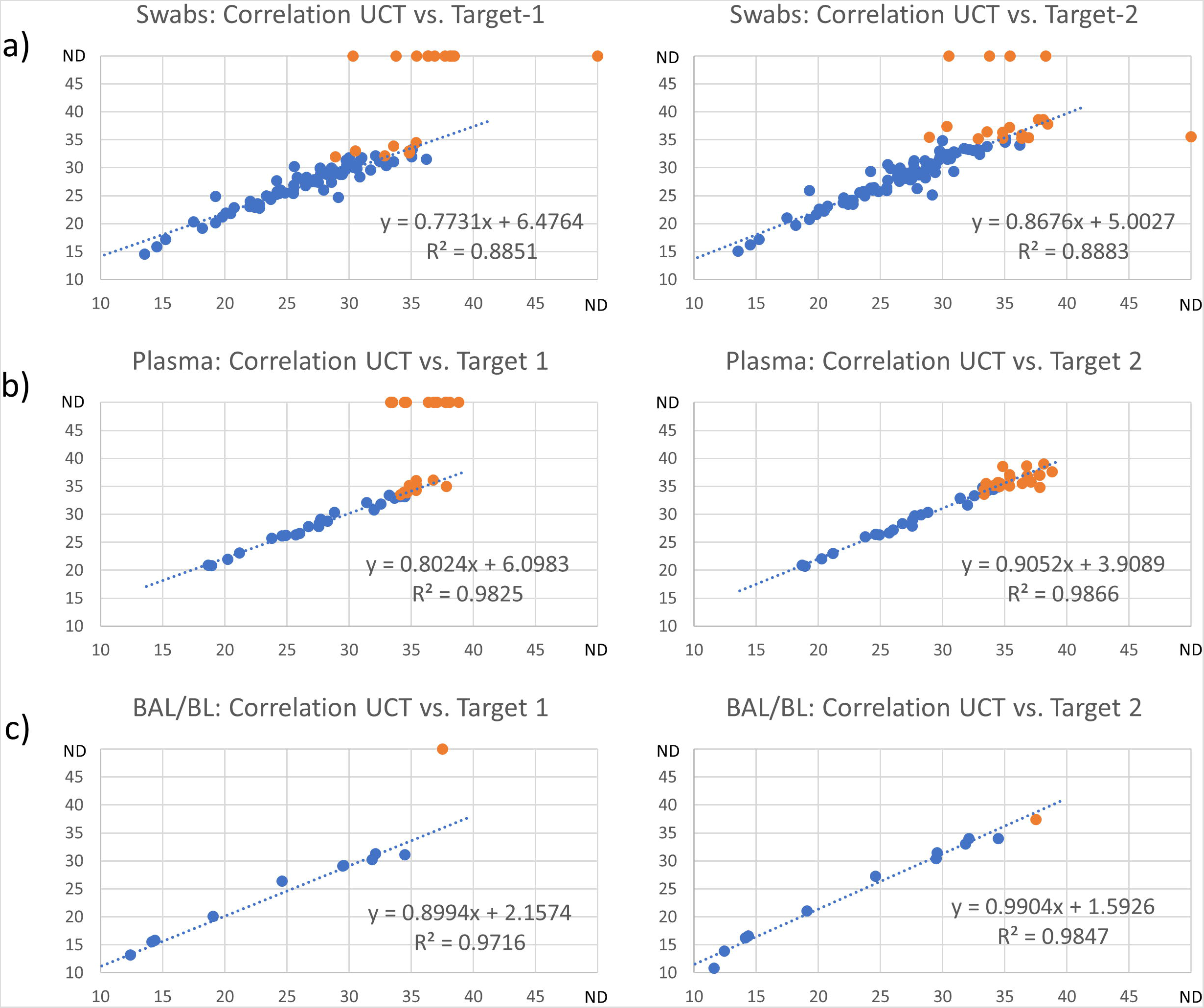
Clinical evaluation of the SARS-CoV-2 IVD compared to the comparator assay, SARS-CoV-2 UCT (utility channel test). Predetermined clinical samples previously analyzed in routine diagnostics were retrieved from storage at –20°C and subjected to testing with the SARS-CoV-2 IVD. X-axis: UCT CT; Y-axis: IVD CT; ND: “Not Detected”; Blue dots: measurements within linear range of Target-2, considered for trendline and correlation; Orange dots: measurements outside linear range, not considered for trendline and correlation.

**Table 1:**
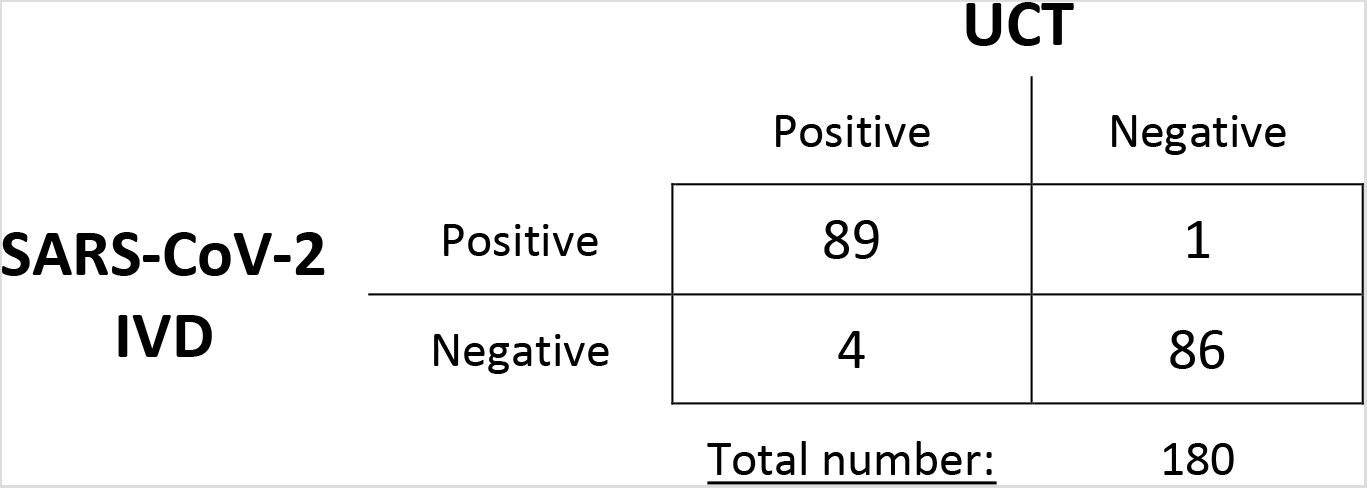
Clinical evaluation of the SARS-CoV-2 IVD assay for 180 predetermined respiratory swab samples. UCT: Utility Channel Test (comparator assay)

All discrepant positive (n = 1) and negative samples (n = 4) were at high CT values (> CT 30), indicating very low concentrations of viral RNA. Concerning CT values, Target-2 (E gene) showed the highest consistency with the reference assay, which is also targeting the viral E gene.

43 predetermined positive plasma samples were correctly identified by the SARS-CoV-2 IVD, though Target-1 failed to detect viral RNA in a number of samples containing only low concentrations (*figure 3b*). For both human plasma and BAL/BL (n = 12, *figure 3c*), results correlated very well with the comparator assay within linear range.

### 4.4 Inter-run variability between different machines and testing sites

A set of samples spiked with SARS-CoV-2 RNA (HH-1) was subjected to testing on a total of four different instruments at three different testing sites. Deviations of CT values did not exceed 1.53 CT in UTM and 0.53 CT in human plasma, indicating excellent comparability of results both between devices and testing sites (*Figure 4*).

**Figure 4:**
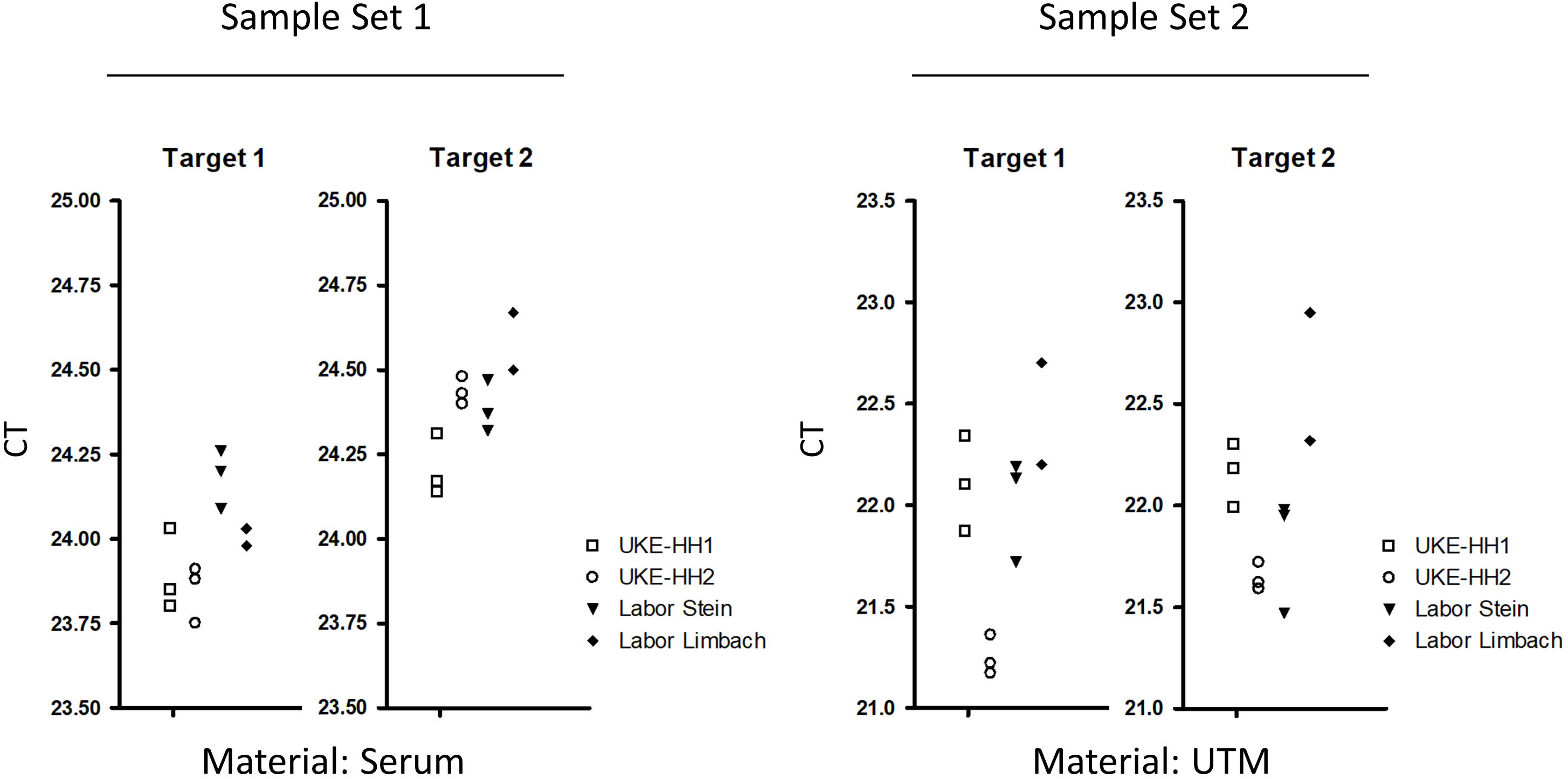
Evaluation of inter-run variability on four different instruments. Sets of seven identical spiked samples were sent to different laboratories in Germany for testing with the SARS-CoV-2 IVD. Material types were as indicated. CT values were reported and compared. UKE-HH1: Instrument No1 at University Medical Center Hamburg-Eppendorf. UKE-HH2: Instrument No2 at University Medical Center Hamburg-Eppendorf. Labor Stein: Moenchengladbach. Labor Limbach: Heidelberg.

## 5 Discussion

The recently released SARS-CoV-2 IVD for the cobas6800/8800 systems made automated high-throughput testing available to a wider range of diagnostic facilities. First studies providing clinical evaluation have demonstrated good agreement with established reference assays (12). The benefits of automation for molecular diagnostics have been discussed extensively in previous studies (11, 14). While a variety of devices offering sample-to-result solutions could already be employed for SARS-CoV-2 testing during the early weeks of the outbreak (including but not limited to the cobas6800 (10), Panther fusion (15) and NeuMoDx 96 (16)), they did require utilization of the systems open modes and the use of lab developed tests (LDT), thus limiting their viability for widespread implementation. The SARS-CoV-2 IVD, however, is a lot more accessible and instruments are relatively widely available in diagnostic labs, thus providing a well standardized common ground for comparable quantification results and viral load monitoring. At present, the assay is limited by relatively narrow recommendations for matrix materials and lack of robust data on analytical and clinical performance. The aim of this study was to go beyond specifications and validate the assay on relevant clinical materials such as BAL/BL and blood, as well as evaluating the efficacy and performance impact of chemical inactivation procedures.

Safety has been a serious concern when handling clinical samples potentially containing SARS-CoV-2 in diagnostic laboratories. While Bio Safety Level 3 (BSL3) conditions are required for procedures that actively enrich or propagate infectious virus (i.e. cell culture), diagnostic samples are usually processed under BSL2 cabinets. Heat inactivation protocols are widespread in some parts of the world as there exists some evidence for their efficacy with SARS-CoV and MERS-CoV (17, 18), however, there are also reports claiming that these procedures may lead to false negative PCR results (19). Pastorino et al. showed that exposure to temperatures as high as 92°C is necessary to completely abolish infectivity, also resulting in substantial degradation of viral RNA in samples (20). Here we show that pre-treating samples by adding 1:1 guanidine hydrochloride solution (cobas PCR Media) reduces plaque formation by at least 3.5 log steps. This procedure did not have significant impact on assay performance in UTM (or eSwab medium). However, guanidine hydrochloride solution alone may not be sufficient to completely inactivate all infectious virus in samples (20, 21).

Since the beginning of the outbreak, some clinical studies have reported on quantitative and semi-quantitative detection of SARS-CoV-2 RNA in different clinical materials over the course of treatment (7, 8, 22). However, quantification results that are precise and reproducible across laboratories require highly standardized reference material, which is not yet available. Vogels et al. recently provided data on performance and linearity for the most commonly used SARS-CoV-2 inhouse assays, demonstrating that linear range and PCR efficiency does vary to a considerable degree (23). Notably, the Sarbeco-E assay by the Charité Berlin (3) and ORF1ab assay by the Hong-Kong University (24) seem to stand out from the crowd, featuring both a maximum of analytical sensitivity and wide linear range. However, other factors such as extraction methods, and choice of enzymes and cyclers will render comparability between sites difficult, even when using the same assays.

In this study we demonstrate that the SARS-CoV-2 IVD dual-target assay is highly efficient and offers a wide linear range in different types of matrix materials outside official specifications. Based on our data, it seems preferable to use the Target-2 (E gene) CT for quantification purposes as it performed slightly better than Target-1 (RdRp gene) in this trial. Compared to the inhouse assay SARS-CoV-2 UCT, the SARS-CoV-2 IVD assay showed good agreement for a total of 180 respiratory swab samples, which is in line with previously reported data (12). Furthermore, good correlation was observed for 43 positive plasma and 12 positive BAL/BL samples within linear range of the IVD assay.

Fully automated RT-PCR systems performing nucleic acid extraction, PCR and signal analysis, provide a standardized common ground between testing sites. In order to evaluate to what degree results are comparable between different instruments, predetermined identical sets of 7 samples were subjected to testing on four different devices at three different testing sites in Germany (UKE in Hamburg, “Labor Limbach” in Heidelberg and “Labor Stein” in Moenchengladbach). There was only minimal variance in CT values for human plasma samples (range dCT: 0.53) and slightly higher in UTM samples (range dCT: 1.53). This further underpins the reproducibility of results obtained from the SARS-CoV-2 IVD which may prove very useful in the context of viral-load monitoring and in clinical multicentre studies.

## 6 Conclusion

In this study we provide evaluation for different matrix materials and clinical performance of the SARS-CoV-2 IVD for the cobas6800/8800 systems. We observed excellent linearity over five to seven log steps in different clinically relevant materials outside assay specifications, including human plasma samples, BAL/BL and swab medium pre-treated for chemical inactivation. Furthermore, deviations in CT values were minimal when identical samples were analyzed at different testing sites. The assay showed excellent correlation with the comparator assay for all specimen types analyzed, indicating compatibility of a variety of clinical materials despite not being officially recommended for use by the manufacturer. Overall, this data demonstrates the viability of the assay for providing reproducible quantification of SARS-CoV-2 RNA in respiratory samples and human plasma for viral load monitoring and clinical studies.

## Data Availability

Available on request.

## 7 Author contribution

ML, SP, LO, MA and NF conceptualized and supervised the study. DN, AF and UE performed the experiments. DN, ML and SP wrote the initial draft of the manuscript. All authors agreed to the publication of the final manuscript.

## 8 Competing interest

UE and ML received speaker honaria and related travel expenses from Roche Diagnostics.

Parts of the material used for this study were provided by Roche Molecular Solutions (Pleasanton, USA).

